## Supplementary material for "Interventions in sexual and reproductive health services addressing violence against women in low- and middle- income countries: a mixed-methods systematic review": An information specialist (AR) applied the search strategy to Medline, Embase, Psycinfo, Cochrane, Cinahl, IMEMR, Web of Science, Popline, Lilacs, WHO

**Supplementary file 1. Search strategy**

Searched 20 August 18
Databases:
(VAW AND Interventions AND LMICs AND healthcare ) less exclusions
See attached Medline strategy
Grey literature:
("violence against women" OR "intimate partner violence" "domestic violence" OR DVA OR IPV OR VAW) AND (intervention* or prevention OR trial*)

**Search results ( databases):**
Medline/Premedline = 1464
Embase= 1403
Psycinfo= 594
Cochrane =61
Cinahl=314
IMEMR =5
Web of Science= 920
Popline= 880
Lilacs= 392
WHO RHL=2
Total= 6035
Total deduplicated =3514

**Search grey literature:**
UNFA=8
SVRI = 5
JPHIEGO =3
USAID =4
WHO (IRIS) SEARO =2
WHO (IRIS) EMRO =3
Google=1
Google Scholar=1
ClinicalTrials.gov =15
WORLD Bank
OTHER= 1
Total= 45
Total deduplicated against database search= 43

**Grand Total ( databases and grey lit) =3557**

Database: Ovid MEDLINE(R) and Epub Ahead of Print, In-Process & Other Non-Indexed Citations and Daily <1946 to August 20, 2018>
Search Strategy:
--------------------------------------------------------------------------------
1 rape/ or domestic violence/ or exp intimate partner violence/ or battered women/ or Gender-Based Violence/ (19658)
2 (violence/ or sex offenses/ or sexual harassment/ or homicide/ or physical abuse/ or coercion/ or crime victims/) and (female/ or women/ or spouses/ or marriage/ or Sexual partners/) (29381)
3 (violence/ or sex offenses/ or sexual harassment/ or homicide/ or physical abuse/ or coercion/ or crime victims/) and (female* or domestic or spous* or partner* or woman or women or married or marriage* or marital or husband* or wife or wives or boyfriend* or girlfriend* or gender-based or non-partner).tw. (13114)

4 ((sexual abuse or sexual harassment or sexual coercion or violent or violence or assault* or beat or beating or batter* or rape* or sex offense* or sexual offense*) adj4 (female* or domestic or spous* or partner* or woman or women or married or marriage* or marital or husband* or wife or wives or boyfriend* or girlfriend* or gender-based or non-partner)).tw. (18139)
5 (IPV or DVA).tw. (5675)
6 (VAW or date rape).tw. (309)
7 ((woman or women) adj3 relationship* adj3 abus*).tw. (90)
8 ((birth control or fertility control or reproductiv* or contraceptiv* or contraception) adj3 (sabotag* or coerc*)).tw. (105)
9 or/1-8 (52401)
10 ((prevent* or intervention* or eliminat* or program* or approach or approaches or trial* or response* or effective or effectiveness or identify or efficacy or what works or outcome* or treatment* or therap* or identification) adj12 (violent or violence or rape* or DVA or IPV or VAW or harassment or sexual offense* or sex offense* or abus* or assault* or beating or beat or coerc* or female* or domestic or spous* or partner* or woman or women or married or marriage* or marital or husband* or wife or wives or boyfriend* or girlfriend* or gender-based or non-partner)).tw. (398870)
11 rape/pc or sex offenses/pc or domestic violence/pc or exp intimate partner violence/pc or battered women/pc or Gender-Based Violence/pc or Sexual Harassment/pc (4812)
12 (psychosocial support or psychological support or education* or training or home visit* or advocacy).tw. (776643)
13 secondary prevention/ or tertiary prevention/ (18314)
14 ((questioning or interviewing or empower*) adj3 (method* or technique*)).tw. (1434)
15 (patient adj3 information).tw. (16715)
16 ((poster* or information or pamphlet* or leaflet*) adj3 (provision or provide*)).tw. (172170)
17 counsel?ing.ti,ab. (81732)
18 exp counseling/ (40659)
19 exp Clinical Trials as Topic/ (316975)
20 exp clinical trial/ (805706)
21 "Controlled Before-After Studies"/ (348)
22 "outcome and process assessment (health care)"/ or "process assessment (health care)"/ (29451)
23 ((program* or process* or service) adj3 evaluation*).tw. (22711)
24 (pretest* or pre-test* or posttest* or post-test* or pre-intervention* or preintervention* or postintervention* or post-intervention*).tw. (52054)
25 (pre* adj12 post*).tw. (579088)
26 ("before and after" or before-after).tw. (243968)
27 or/10-26 (2980515)
28 9 and 27 (24093)
29 Developing Countries/ (70541)
30 (developing countr* or emerging econom* or third world).tw. (55717)
31 ((low or middle) adj4 income countr*).ti,ab. (16763)
32 LMIC*.ti,ab. (2703)
33 (Afghanistan or Benin or Burkina Faso or Burundi or Cambodia or Central Africa or Chad or Comoros or Congo or Eritrea or Ethiopia or Gambia or Guinea or Bissau or Haiti or North Korea or Liberia or Madagascar or Malawi or Mali or Mozambique or Nepal or Niger or Rwanda or Sierra Leone or Somalia or Tanzania or Togo or Uganda or Zimbabwe).mp. (289340)
34 (Armenia or Bangladesh or Bhutan or Bolivia or Cabo Verde or Cameroon or Cote d'Ivoire or Ivory Coast or Djibouti or Egypt or El Salvador or Georgia or Ghana or Guatemala or Guyana or Honduras or India or Indonesia or Kenya or Kiribati or Kosovo or Kyrgyz* or Lao or Laos or Lesotho or Mauritania or Micronesia or Moldova or Mongolia or Morocco or Myanmar or Nicaragua or Nigeria or Pakistan or Philippines or Samoa or Sao Tome or

Principe or Senegal or Solomon Islands or Sri Lanka or Sudan or Swaziland or Syria* or Tajikistan or Timor Leste or Ukraine or Uzbekistan or Vanuatu or Vietnam or West Bank or Gaza or Yemen or Zambia).mp. (392622)
35 (Albania or Angola or Argentina or Panama or Tunisia or Palau or Tunisia or Herzegovina or Fiji or Namibia or Algeria or Gabon or Nauru or Grenada or Paraguay or Peru or Azerbaijan or Grenadines or Romania or Belarus or Iran or Russia* or Belize or Iraq or Bosnia or Jamaica or Serbia).mp. (189750)
36 (Botswana or Jordan or South Africa or Brazil or Kazakhstan or Saint Lucia or St Lucia or Bulgaria or Lebanon or Saint Vincent or St Vincent or China or Libya or Suriname or Colombia or Macedonia or Thailand or Costa Rica or Malaysia or Tonga).mp. (435209)
37 (Cuba or Maldives or Turkey or Dominica* or Marshall Islands or Turkmenistan or Mauritius or Tuvalu or Mexico or Venezuela or Ecuador or Montenegro).mp. (122538)
38 or/29-37 (1403313)
39 28 and 38 (4702)
40 exp maternal health services/ or exp reproductive health services/ or family planning services/ (67984)
41 exp pregnancy/ or exp pregnancy trimesters/ or pregnant women/ or peripartum period/ or exp pregnancy complications/ or exp fetal therapies/ or exp Obstetric surgical procedures/ or exp postpartum period/ or obstetric nursing/ or midwifery/ (914606)
42 exp maternal-child nursing/ (5549)
43 (adolescent health services/ or community mental health services/ or community health services/ or rural health services/ or rural nursing/ or family health/ or adolescent health/ or exp primary health care/ or exp general practice/ or general practitioners/ or physicians, family/) and (women or woman or reproductive or sexual health* or "STI" or STD* or "STIS" or contracept* or abortion or childbirth or pregnan*).mp. (26039)
44 reproductive medicine/ or gynecology/ or obstetrics/ or "Obstetrics and Gynecology Department, Hospital"/ (35849)
45 ((sexual or reproductive) adj3 (education or healthcare or care or service* or program* or clinic*)).mp. (12944)
46 ((sexual or reproductive) adj3 (education or health* or care)).jn,in. (14583)
47 ((pregnan* or birth or childbirth or midwife* or midwive* or " mother and baby" or obstetric* or maternal or maternity or postpartum or antepartum or postnatal or post-natal or ante-natal or antenatal or prenatal or pre-natal or perinatal or peri-natal or contraception or contraceptiv* or abortion or fertility or gynae* or gyne* or STD* or "STI" or "STIS" or sexually transmitted or PMS or premenstrual syndrome) adj3 (care or healthcare or clinic* or service* or treatment*)).tw. (109386)
48 (cervical adj2 (smear* or screening)).tw. (11930)
49 vaginal smears/ or papanicolaou test/ (22149)
50 exp Sexually Transmitted Diseases/di, pc, rh, th (103211)
51 exp Women's Health/ (26647)
52 exp Menstruation Disturbances/di, pc, rh, th (5369)
53 ((woman* or women*) adj3 health*).jn,in,mp. (106372)
54 family planning*.jn,mp,in. (50733)
55 exp "diagnostic techniques, obstetrical and gynecological"/ (124519)
56 reproductive health/ or sexual health/ (2688)
57 or/40-56 (1238548)
58 39 and 57 (1822)
59 letter/ (997536)
60 editorial/ (466054)
61 news/ (190965)
62 exp historical article/ (382354)
63 Anecdotes as topic/ (4721)
64 comment/ (730950)
65 (letter or editorial or comment*).ti. (160815)
66 exp animals/ not humans/ (4489180)

67 exp Animals, Laboratory/ (820540)
68 exp Animal Experimentation/ (8786)
69 exp Models, Animal/ (516652)
70 exp rodentia/ (3047766)
71 (rat or rats or mouse or mice or rodent* or animal*).ti. (1394667)
72 or/59-71 (7516664)
73 58 not 72 (1776)
74 limit 73 to yr="2005 -Current" (1464)
