## Supplementary material for "Interventions in sexual and reproductive health services addressing violence against women in low- and middle- income countries: a mixed-methods systematic review": Searches identified 6082 records, we assessed 313 full text reports and included 32 reporting on 26 studies (Figu

### Supplementary file 2. Characteristics of included studies by intervention category and level of evidence

| Study, publication | Country | Setting | Sample characteristics | Intervention (n) vs comparison (n) | Design/ | Follow up | Primary outcomes of interest for this review | Secondary outcomes of interest for this review |
|---|---|---|---|---|---|---|---|---|
| **Response to VAW during routine SRH consultation (n=10)** | | | | | | | | |
| Vakily 2017[58] | Iran | Antenatal clinic, 32 outpatient health centres | HCPs (midwives) | 2-hour HCP training computer assisted (35) vs face-to-face (35) | RCT | 2 months | | Knowledge and attitudes about DV |
| Brown 2018[32] | South Africa | HIV testing and counselling, community, NGO | HIV positive women 18+ | 7-minute integrated HIV-IPV consultation over phone (166) vs standard care (83) | RCT | 1 month | IPV upon partner notification of serostatus, harm | Perceived safety, safety behaviours, access to HIV treatment |
| *Haberland 2016[38] | Kenya | HIV testing in antenatal clinic, hospital with GBV centre | HCPs (HIV testing counsellors) Pregnant women 15-49 | HCP training and ongoing support, 29-minute integrated HIV-IPV consultation, referral to IPV counsellor in ANC clinic (337) vs standard care (351) | RCT Nested mixed-method process evaluation | 1 month | Any IPV, harm | IPV screening, referrals Women's knowledge, attitudes, self-esteem, perceived intervention effect, HIV care Intervention acceptability |
| Abeid 2016[28] | Tanzania | Post-rape care service, 5 health centres and referral hospitals | HCPs (doctors, nurses, assistant medical/clinical officers) | 5-day training, guidelines, infrastructure improvement (100) vs minimal intervention (53) | Controlled before-after | 12 months | | Knowledge and attitudes about sexual violence and post-rape care Provision of post-rape care |
| Jayatilleke 2015[39] | Sri Lanka | Antenatal clinic, community | HCPs (midwives) | 4-day training, handbook, external referral (408) | Uncontrolled before-after | 6 months | | Knowledge, practices, responsibility, readiness for identifying and responding to IPV, provision of referrals |
| Matseke 2013[46] | South Africa | HIV testing and counselling in antenatal clinic, 16 primary health care clinics | Pregnant women 18+ | HPC training, 30-minute integrated ANC-IPV consultation, external referral (160) | Uncontrolled before-after | 3 months | Perceived risk of becoming a victim of femicide | |
| Smith 2013[54] | Kenya, Ethiopia, Jordan, Democratic Republic of Congo | Post-rape care service, 35 humanitarian settings, NGO | HCPs (doctors, nurses, midwives) | 4-day training, infrastructure improvement (106) | Uncontrolled before-after Qualitative study | 3 months | | Attitudes, knowledge, skills on sexual violence and post-rape care, provision of post-rape care |
| Laisser 2011[45] | Tanzania | Antenatal clinic, hospital | HCPs (clinical/medical | HCP training (39), infrastructure improvement, integrated ANC-IPV | Cross sectional | 3 weeks | | Intervention acceptability |

| Study, publication | Country | Setting | Sample characteristics | Intervention (n) vs comparison (n) | Design/ | Follow up | Primary outcomes of interest for this review | Secondary outcomes of interest for this review |
|---|---|---|---|---|---|---|---|---|
| | | | officers, nursing officers)<br>Women 18+ | consultation (102), external referral | Qualitative study | | | |
| Undie 2016[57] | Kenya | HIV testing in antenatal clinic, hospital with GBV centre | Women | HCP training, integrated HIV-IPV consultation (1210), assisted referral to on-site GBV centre | Cross sectional<br>Qualitative study | 7 months | | IPV screening, referrals<br>Intervention acceptability |
| Cristofides 2010[33] | South Africa | HIV testing and counselling, primary health care clinic | HCPs (lay counsellors)<br>Women | HCP training (16), integrated HIV-IPV consultation (35), external referral | Qualitative study | 2 weeks | | Intervention acceptability |
| **Response to VAW during routine SRH consultation plus community engagement (n=9)** | | | | | | | | |
| Cockcroft 2019[34] | Nigeria | Universal home visits, 4 communities | Pregnant women 14-49 | HCP training, infrastructure improvement, integrated DV-universal home visits that discussed domestic violence, heavy work in pregnancy, ignorance of danger signs, and lack of spousal communication with pregnant women (1837) and their partners vs delayed intervention (1853) | Cluster RCT | 12 months | Physical DV, pregnancy delivery, postnatal complications | Use of SRH services |
| Settergren 2018[60] | Tanzania | HIV/AIDS services, hospital, and health centre | Women 15-49 | Systems level activities, HCP training, infrastructure improvement, integrated-HIV-GBV consultation, onsite and external referral, community, and couple education (6 facilities, 656 women) vs standard care (6 facilities, 643 women) | Cluster RCT | 28 months | Any IPV | Provision of services to IPV positive patients |
| Wagman 2015[59] | Uganda | HIV testing and counselling, community | Women 15-49 | HCP training, integrated HIV-IPV consultation, onsite referral (6 facilities, 1812 women) vs standard care (5 facilities, 2127 women) | Cluster RCT | 4 years and 7 months | Physical, emotional, sexual IPV, HIV incidence | Risk behaviours and HIV disclosure |
| *Bott 2014[30 37 48] | Dominican Republic, Peru, Venezuela | 3 family planning clinics, NGO | HCP (doctors, nurses, midwives, counsellors, social workers, psychologists, receptionists)<br>Women 12+ | Systems level activities, HCP training and ongoing support, infrastructure improvement, integrated GBV-family planning consultation, referral to onsite GBV specialist (4 clinics) | Uncontrolled before-after<br>Qualitative study | 3 years | | HCPs attitudes, knowledge, readiness for identifying and responding to GBV<br>Intervention acceptability |

| Study, publication | Country | Setting | Sample characteristics | Intervention (n) vs comparison (n) | Design/ | Follow up | Primary outcomes of interest for this review | Secondary outcomes of interest for this review |
|---|---|---|---|---|---|---|---|---|
| Kim 2007[42 43] | South Africa | Post-rape care service, hospital | Survivors of sexual violence | Systems level activities, 2-day HCP training (334), infrastructure improvement, community education on post-rape care | Uncontrolled before-after | No information | | Use, quality, and cost of post-rape care service |
| Bress 2018[31] | Democratic Republic of Congo | Post-rape care service, 12 primary care clinics and referral hospital | Survivors of sexual violence 12+ | HCP training and ongoing support, infrastructure improvement, community education on post-rape care (13 sites, 2081 survivors) | Cross-sectional | 4 years | | Provision of post-rape kit |
| Samandari 2016[36 49] | Guinea | Family planning clinic | HCPs (nurse, midwife, counsellor, support/admin staff) Women | System level activities, 7-day HCP training and ongoing support (4), integrated family planning-IPV consultation (171), external referral, community education | Cross-sectional Qualitative study | 4 months | | IPV identification, safety planning, referrals Intervention acceptability |
| Sithole 2018[53] | Zimbabwe | Comprehensive post-rape care service, 8 policlinics, NGO | HCPs (doctors, nurses, managers) Survivors of sexual violence | HCP training (80), infrastructure improvement, post-rape care (1669), community education on post-rape care | Cross sectional service evaluation | 4 years | | HCPs knowledge about post-rape care Provision of post-rape care |
| Turan 2013[56] | Kenya | Antenatal clinic, primary health care clinic | HCPs (all clinic staff including admin, community volunteers, lay health workers) Pregnant women | 40-hour HCP training, integrated ANC-GBV consultation (134), assisted external referral, community education | Cross sectional Qualitative study | 5 months | | GBV identification, referrals Intervention acceptability |
| **Response to VAW in addition to routine SRH consultation (n=7)** | | | | | | | | |
| Cripe 2010[35] | Peru | Antenatal clinic, hospital | Pregnant women 18-45 with IPV experience | 1* 30-minute psychosocial counselling session by social worker, resource card, external referral (110) vs minimal intervention (110) | RCT | Prenatal appointment to 1 week after delivery | Quality of life | Safety behaviours, use of community resources |
| Khalili 2019[40 41] | Iran | Antenatal clinic, University health centers | Pregnant women 20+ with IPV experience | 4*90-minute psychoeducational sessions by counsellor (50) vs standard care (50) | RCT | 2 months | Verbal and physical IPV, psychological distress | |
| Mutisya 2018[47] | Kenya | Antenatal clinic, 12 primary health care clinics | Pregnant women 18-45 with IPV experience | 1-3*30-35-minute psychosocial counselling sessions by researcher, risk assessment, safety planning, resource card, external referral | RCT | 6 months | Physical, emotional, severe combined IPV and harassment, depression | |

| Study, publication | Country | Setting | Sample characteristics | Intervention (n) vs comparison (n) | Design/ | Follow up | Primary outcomes of interest for this review | Secondary outcomes of interest for this review |
|---|---|---|---|---|---|---|---|---|
| | | | | (141) vs minimal intervention (142) | | | | |
| Sapkota 2020[50 51] | Nepal | Antenatal clinic, hospital | Pregnant women 18+ with DV experience | 1*35-45-minute psychosocial session by counsellor, resource card, contact details of the counsellor (70) vs minimal intervention (70) | RCT<br>Nested qualitative study | Prenatal appointment to 6 weeks after delivery | Depression, anxiety, quality of life | Self-efficacy, safety behaviours, social support<br>Intervention acceptability |
| Sikkema 2018[44 52] | South Africa | HIV testing and treatment, primary health care clinic | HIV positive women 18+ with experience of sexual violence | 4 individual and 3 group*90-minute psychosocial training sessions by trained lay provider (32) vs standard care (32) | RCT<br>Nested qualitative study | 6 months | PTSD | Coping strategies, engagement with HIV treatment<br>Intervention acceptability |
| Tanghizaden 2018[55] | Iran | Antenatal clinic, 16 health centres | Pregnant women with IPV experience | 4*90-minute psychosocial training sessions on problem-solving skills by researcher (125) vs standard care (132) | RCT | 3 months | Physical, psychological, sexual IPV | |
| Arora 2019[29] | India | Antenatal clinic, 2 hospitals | Pregnant women with DV experience | ≥2*30-45-minute psychosocial sessions by counsellor (155) | Uncontrolled before-after | First prenatal appointment to 6 weeks after delivery | Physical, emotional, financial DV, physical health, emotional health | Knowledge and attitudes about DV, coping behaviours |

Note. * grey literature. NGO non-governmental (third sector) organisation. RCT randomised controlled trial. HCP health care providers. IPV intimate partner violence. DV domestic violence. GBV gender-based violence. HIV human immunodeficiency viruses. PTSD posttraumatic stress disorder. SRH sexual and reproductive
