## Supplementary material for "Interventions in sexual and reproductive health services addressing violence against women in low- and middle- income countries: a mixed-methods systematic review": Most quantitative studies were at high risk of bias

**Online supplementary file 3. Quality appraisal**

### Risk of bias in randomised controlled trials

| Study ID | Bias arising from the randomization process | Bias due to deviations from the intended interventions (assignment) | Bias due to deviations from the intended interventions (adherence) | Bias due to missing outcome data | Bias in measurement of the outcome | Bias in selection of the reported result | Overall risk of bias |
|---|---|---|---|---|---|---|---|
| Brown 2018[32] | Low | Some concerns | High | Low | Low | Low | High |
| Cockcroft 2019[34] | Low | Low | Low | Low | Low | Low | Low |
| Cripe 2013[35] | Low | Low | Low | Low | Some concerns | Some concerns | Some concerns |
| Haberland 2016[38] | Some concerns | Low | High | Low | Some concerns | Some concerns | High |
| Khalili 2020[40] | Low | High | High | Low | High | Low | High |
| Mutisya 2018[47] | Low | High | High | Low | High | Some concerns | High |
| Sapkota 2020[50] | Low | Low | Low | Low | Low | Low | Low |
| Settergren 2018[60] | Low | Low | High | Low | High | Low | High |
| Sikkema 2018[52] | Low | Low | Low | High | Some concerns | Some concerns | High |
| Tanghizadeh 2018[55] | Low | Low | Low | Low | Low | Some concerns | Some concerns |
| Vakily 2017[58] | Some concerns | Some concerns | High | High | Low | Some concerns | High |
| Wagman 2015[59] | Some concerns | High | Some concerns | Low | High | Some concerns | High |

### Risk of bias (EPOC criteria) in controlled before-after studies

| Study ID | Was the allocation sequence adequately generated? | Was the allocation adequately concealed? | Were baseline outcome measurements similar? | Were baseline characteristics similar? | Were incomplete outcome data adequately addressed? | Was knowledge of the allocated interventions adequately prevented during the study? | Was the study adequately protected against contamination ? | Was the study free from selective outcome reporting? | Was the study free from other sources of bias? | Overall risk of bias |
|---|---|---|---|---|---|---|---|---|---|---|
| Abeid 2016[28] | No | No | Yes | Yes | Yes | Unclear | Unclear | Yes | Unclear | High |

### Risk of bias (EPOC criteria) in studies without a control group

| Study ID | Was the intervention independent of other changes? | Was the shape of the intervention effect pre-specified? | Was the intervention unlikely to affect data collection? | Was knowledge of the allocated interventions adequately prevented during the study? | Were incomplete outcome data adequately addressed? | Was the study free from selective outcome reporting? | Was the study free from other risk of bias? | Overall risk of bias |
|---|---|---|---|---|---|---|---|---|
| Arora 2019[29] | No | Yes | Yes | No | Yes | No | Unclear | High |
| Bott 2004[30] | Unclear | Unclear | Yes | No | No | Unclear | Unclear | High |
| Bress 2019[31] | No | No | No | No | Unclear | Yes | Unclear | High |

| Study ID | Was the intervention independent of other changes? | Was the shape of the intervention effect pre-specified? | Was the intervention unlikely to affect data collection? | Was knowledge of the allocated interventions adequately prevented during the study? | Were incomplete outcome data adequately addressed? | Was the study free from selective outcome reporting? | Was the study free from other risk of bias? | Overall risk of bias |
|---|---|---|---|---|---|---|---|---|
| Jayatilleke 2015[39] | Unclear | Yes | Yes | No | Unclear | Yes | Unclear | High |
| Kim 2007[42] | No | Yes | Yes | No | Unclear | No | No | High |
| Laisser 2011[45] | No | No | No | No | Unclear | Yes | Unclear | High |
| Matseke 2013[46] | Unclear | Yes | Yes | No | No | Yes | No | High |
| Samandari 2016[49] | No | Yes | Yes | No | Yes | Yes | Unclear | High |
| Sithole 2018[53] | Unclear | No | No | No | Unclear | No | Unclear | High |
| Smith 2013[54] | No | Yes | Yes | Unclear | Yes | Yes | No | High |
| Turan 2013[56] | Unclear | No | No | No | Unclear | Yes | Unclear | High |
| Undie 2016[57] | No | No | No | No | Unclear | Unclear | Unclear | High |

### Quality appraisal of qualitative studies

| CASP signalling questions | Bott 2004[30] | Christofides 2010[33] | Haberland 2016[38] | Laisser 2011[45] | Samandari 2016[49] | Sapkota 2020[51] | Sikkema 2018[44] | Smith 2013[54] | Turan 2013[56] | Undie 2016[57] |
|---|---|---|---|---|---|---|---|---|---|---|
| 1. Interprets subjective experiences? | Yes | Yes | Yes | Yes | Yes | Yes | Yes | Yes | Yes | Yes |
| 2. Right methodology? | Yes | Yes | Yes | Yes | Yes | Yes | Yes | Yes | Yes | Yes |
| 3. Appropriate design? | Yes | Yes | Yes | Yes | Yes | Yes | Yes | Yes | Yes | Yes |
| 4. Design justified? | Yes | No | No | Yes | No | Yes | Yes | No | No | No |
| 5. Ethical issues considered? | No | No | Yes | Yes | Yes | Yes | Yes | Yes | Yes | Yes |
| 6. Credibility established? | Yes | No | No | Yes | Yes | Yes | Yes | Yes | Yes | Yes |
| 7. Transferability established? | No | Yes | No | Yes | Yes | Yes | Yes | Yes | No | No |
| 8. Purpose established? | Yes | Yes | Yes | Yes | Yes | Yes | Yes | Yes | No | Yes |
| 9. Recruitment appropriate? | Yes | Yes | Yes | Yes | Yes | Yes | Yes | Yes | Yes | Yes |
| 10. Selection of participants explained? | No | Yes | Yes | Yes | Yes | Yes | Yes | No | Yes | No |
| 11. Participants appropriate? | No | No | No | Yes | Yes | No | Yes | Yes | Yes | No |
| 12. Discussed recruitment? | No | Yes | No | No | No | No | Yes | No | No | No |
| 13. Justified setting? | Yes | Yes | Yes | Yes | Yes | Yes | Yes | Yes | Yes | No |
| 14. How data were collected? | Yes | Yes | Yes | Yes | Yes | Yes | Yes | Yes | Yes | Yes |
| 15. Justified data collection method? | No | No | Unsure | Yes | No | Yes | Yes | No | No | No |

| CASP signalling questions | Bott 2004[30] | Christofides 2010[33] | Haberland 2016[38] | Laisser 2011[45] | Samandari 2016[49] | Sapkota 2020[51] | Sikkema 2018[44] | Smith 2013[54] | Turan 2013[56] | Undie 2016[57] |
|---|---|---|---|---|---|---|---|---|---|---|
| 16. Described data collection method? | No | Yes | Yes | Yes | Yes | Yes | Yes | Yes | Yes | Yes |
| 17. Form of data clear? | No | Yes | Yes | Yes | Yes | Yes | Yes | Yes | Yes | Yes |
| 18. Described how data were reduced/transformed for analysis? | No | No | No | Yes | Yes | Yes | Yes | Yes | Yes | Yes |
| 19. Discussed interpretation of findings? | Yes | No | No | Yes | Yes | Yes | Yes | No | Yes | Yes |
| 20. Ensured neutrality? | No | No | No | No | Yes | Yes | Yes | No | Yes | No |
| **Total (Yes/No/Unsure)** | **10/10/0** | **12/8/0** | **11/8/1** | **18/2/0** | **17/3/0** | **18/2/0** | **20/0/0** | **14/6/0** | **15/5/0** | **12/8/0** |
