## Supplemental Data 1 for "Interventions in sexual and reproductive health services addressing violence against women in low- and middle- income countries: a mixed-methods systematic review"

### Supplementary file 4. Effects and outcomes of interventions on response to VAW during routine SRH consultation

| Outcomes | Results (95% CI, p) | Effect (95% CI) | N of participants (design/follow up) | Study | Risk of bias | Direction of effect/interpretation |
|---|---|---|---|---|---|---|
| **Direct effect on health-related cognition and emotions** | | | | | | |
| **HCP knowledge about VAW, relevant procedures (n=4)** | | | | | | |
| Mean (SD) score of knowledge about domestic violence, post-intervention | Group training = 16.1 (1.9)<br>CD training = 17.7 (1.1)<br>p<0.001 | - | 35 HCPs (RCT/2 month) | Vakily 2017[58] | High | 2-hour CD training improved HCP knowledge about domestic violence more than group training |
| Change in proportion with correct knowledge on sexual violence | Intervention = 31.4%<br>Control = -22.3% | Net effect = 53.7% (32.2; 75.1) | HCPs (CBA/12 months | Abeid 2016[28] | High | 5-day training, guidelines, infrastructure improvement improved HCPs knowledge about sexual violence |
| Median (IQR) score of knowledge about IPV | Pre-intervention = 0.62 (0.43-0.81)<br>Post-intervention = 0.88 (0.82-0.94)<br>p<0.001 | - | 408 HCPs (UBA/6 months) | Jayatilleke 2015[39] | High | 4-day training, handbook, external referral improved HCP knowledge about IPV |
| Mean (95% CI) score of knowledge in providing care to sexual assault survivors | Pre-intervention = 49.09 (45.57; 51.34)<br>Post-intervention = 61.59 (59.04; 64.42) | MD = 12.50 (10.29; 16.24) | 106 HCPs (UBA/3 months) | Smith 2013[54] | High | 4-day training, infrastructure improvement contributed towards improved HCP knowledge about providing clinical care to survivors of sexual violence |
| **HCP attitudes about VAW (n=3)** | | | | | | |
| Mean (SD) score of attitudes about domestic violence, post-intervention | Group training = 46.9 (4.9)<br>CD training = 45.4 (6.4)<br>p = 0.3 | - | 35 HCPs (RCT/2 month) | Vakily 2017[58] | High | Neither group nor CD 2-hour training had effect on HCP attitudes about domestic violence |
| Mean (95% CI) score of attitudes about sexual violence | Pre-intervention = 71.76 (66.79; 73.14)<br>Post-intervention = 77.20 (72.53; 78.34) | MD = 5.44 (1.89; 8.98) | 106 HCPs (UBA/3 months) | Smith 2013[54] | High | 4-day training, infrastructure improvement contributed towards improved attitudes about sexual violence |
| Change in proportion with accepting attitude towards sexual violence | Intervention = -4.1%<br>Control = 6.8% | Net effect = -10.9% (-27.2; 5.5) | HCPs (CBA/12 months | Abeid 2016[28] | High | 5-day training, guidelines, infrastructure improvement had no effect on HCP attitudes about sexual violence |
| **HCP readiness for identifying and responding to VAW (n=2)** | | | | | | |
| Median (IQR) score of perceived barriers to IPV identification and response | Pre-intervention = 2.43 (2.14-3.14)<br>Post-intervention = 1.14 (1.14-1.28)<br>p<0.001 | | 408 HCPs (UBA/6 months) | Jayatilleke 2015[39] | High | 4-day training, handbook, external referral reduced HCP perceived barriers to identifying and responding to IPV |
| Median (IQR) score of perceived responsibilities to identify and respond to IPV | Pre-intervention = 3.20 (2.80-3.95)<br>Post-intervention = 4.60 (4.20-4.80)<br>p<0.001 | | | | | 4-day training, handbook increased HCP perceived responsibility and self-confidence to identify and respond to IPV |

| Outcomes | Results (95% CI, p) | Effect (95% CI) | N of participants (design/follow up) | Study | Risk of bias | Direction of effect/interpretation |
|---|---|---|---|---|---|---|
| Median (IQR) score of self-confidence to identify and respond to IPV | Pre-intervention = 1.81 (1.38-2.12)<br>Post-intervention = 2.75 (2.62-2.88)<br>p<0.001 | | | | | |
| Mean (95% CI) score of HCPs' confidence in providing care to sexual assault survivors | Pre-intervention = 58.16 (53.86; 63.90)<br>Post-intervention = 72.66 (66.21; 74.30) | MD = 14.50 (8.22; 20.77) | 106 HCPs (UBA/3 months) | Smith 2013[54] | High | 4-day training, infrastructure improvement contributed towards improved confidence in providing clinical care to sexual assault survivors |
| **Women knowledge about VAW (n=1)** | | | | | | |
| Mean difference (95% CI) in women's IPV knowledge score, post-intervention | - | MD = 0.16<br>Crude β=0.176 (0.02; 0.033)<br>Adjusted β=0.155 (0.00-0.31) | 337 women (RCT/1 month) | Haberland 2016[38] | High | HCP training and ongoing support, 29-minute integrated HIV-IPV consultation, referral to IPV counsellor in ANC clinic improved knowledge about IPV and women's rights among pregnant women |
| Mean (SD) score of learning about women's rights in relationship | Intervention = 2.6 (1.1)<br>Control = 2.0 (1.0)<br>p<0.0001 | - | | | | |
| **Women attitudes about VAW (n=1)** | | | | | | |
| Proportion (n) who justified wife beating, post-intervention | Intervention = 18.3% (49/268)<br>Control = 21.8% (58/267)<br>p=0.33 | - | 337 women (RCT/1 month) | Haberlan 2016[38] | High | HCP training and ongoing support, 29-minute integrated HIV-IPV consultation, assisted onsite referral had no effect on attitudes about IPV among pregnant women |
| **Women readiness for addressing VAW (n=1)** | | | | | | |
| Proportion (n) who felt more confident in how deserve to be treated, post-intervention | Intervention = 82% (73/107)<br>Control = 71.6% (73/134)<br>p=0.12 | - | 337 women (RCT/1 month) | Haberlan 2016[38] | High | HCP training and ongoing support, 29-minute integrated HIV-IPV consultation, assisted onsite referral had no effect on self-confidence among pregnant women |
| **Intermediate effects on health-related behaviour and practices** | | | | | | |
| **VAW enquiry rate (n=2)** | | | | | | |
| Proportion (n) screened for IPV, post-intervention | Intervention = 76% (81/107)<br>Control = 22% (29/134)<br>p<0.0001 | - | 337 women (RCT/1 month) | Haberland 2016[38] | High | HCP training and ongoing support, 29-minute integrated HIV-IPV consultation, assisted onsite referral increased IPV enquiry rate |
| Proportion (n) who discussed IPV | Pre-intervention = 67.3% (201)<br>Post-intervention = 96.5% (387)<br>p<0.01 | - | 408 HCPs (UBA/6 months) | Jayatilleke 2015[39] | High | 4-day HCP training, handbook, external referral increased IPV enquiry rate |
| **Provision of referrals to VAW services (n=2)** | | | | | | |
| Proportion (n) referred to GBV centre of those disclosed, post-intervention | Intervention = 56% (19/34)<br>Control = 33% (3/9)<br>p=0.28 | - | 337 women (RCT/1 month) | Haberland 2016[38] | High | HCP training and ongoing support, 29-minute integrated HIV-IPV consultation, referral to IPV counsellor in ANC clinic had no effect on referral rate |

| Outcomes | Results (95% CI, p) | Effect (95% CI) | N of participants (design/follow up) | Study | Risk of bias | Direction of effect/interpretation |
|---|---|---|---|---|---|---|
| Proportion (n) referred to the medical officer or Heath/IPV services | Pre-intervention = 6.5% (13)<br>Post-intervention = 22.4% (87)<br>p not reported | - | 408 HCPs (UBA/6 months) | Jayatilleke 2015[39] | High | 4-day training, handbook, external referral had no effect on referral rates to external IPV services |
| Proportion (n) referred to GBV centre of those disclosed, post-intervention | 75% (73/95) | - | 1210 women (Cross sectional/7 months) | Undie 2016[57] | High | HCP training, integrated IPV-HIV consultation, assisted referral contributed towards 75% referral rate to on-site GBV centre |
| **Provision of post-rape care (n=2)** | | | | | | |
| Change in proportion who used a rape kit | Intervention = 59.6%<br>Control = -4.5% | Net effect = 64.1% (46.7; 81.5) | 100 HCPs (CBA/12 months) | Abeid 2016[28] | High | 5-day training, guidelines, infrastructure improvement contributed towards improvement on 10 out of 18 indicators of post-rape care |
| Change in proportion who gave prophylactic treatment for STI | Intervention = 10.9%<br>Control = 3.4% | Net effect 7.5% = (-14.5; 29.5) | | | | |
| Proportion of eligible patients who received emergency contraception | Pre-intervention = 50%<br>Post-intervention = 82%<br>p<0.01 | - | 60 patients (UBA/3 months) | Smith 2013[54] | High | 4-day training, infrastructure improvement contributed towards improvement on 6 out of 10 indicators of post-rape care service |
| Proportion of eligible patients who received HIV post-exposure prophylaxis | Pre-intervention = 42%<br>Post-intervention = 92%<br>p<0.001 | - | | | | |
| Proportion of eligible patients who received STI prophylaxis and treatment | Pre-intervention = 45%<br>Post-intervention = 96%<br>p<0.01 | - | | | | |
| **VAW disclosure rate (n=4)** | | | | | | |
| Proportion (n) who disclosed IPV of those screened, post-intervention | Intervention = 32% (34/107)<br>Control = 7% (9/134)<br>p<0.0001 | - | 337 women (RCT/1 month) | Haberland 2016[38] | High | HCP training and ongoing support, 29-minute integrated HIV-IPV consultation, referral to IPV counsellor in ANC clinic increased IPV identification rate |
| Proportion (n) who identified at least one IPV during past 3 months | Pre-intervention = 73.3% (299)<br>Post-intervention = 98.5% (402)<br>p<0.001 | - | 408 HCPs (UBA/6 months) | Jayatilleke 2015[39] | High | 4-day training, handbook increased IPV identification rate |
| Proportion who disclosed IPV of those screened | 62% | - | 102 women (Cross-sectional/3 weeks) | Laisser 2011[45] | High | HCP training, infrastructure improvement, integrated ANC-IPV consultation, external referral contributed towards 62% IPV identification rate |
| Proportion (n) who disclosed IPV of those screened | 8% (95/1210) | - | 1210 women (Cross-sectional/7 months) | Undie 2016[57] | High | HCP training, integrated HIV-IPV consultation, assisted onsite referral contributed towards 8% IPV identification rate |
| **VAW referral uptake (n=2)** | | | | | | |
| Proportion (n) who used GBV centre out of those referred, post-intervention | Intervention = 63% (12/19)<br>Control = 100% (3/3)<br>p=0.52 | - | 337 women (RCT/1 month) | Haberland 2016[38] | High | HCP training and ongoing support, 29-minute integrated HIV-IPV consultation, referral to IPV |

| Outcomes | Results (95% CI, p) | Effect (95% CI) | N of participants (design/follow up) | Study | Risk of bias | Direction of effect/interpretation |
|---|---|---|---|---|---|---|
| | | | | | | counsellor in ANC clinic had no effect on uptake of referrals to on-site GBV centre |
| Proportion (n) who used GBV centre out of those referred, post-intervention | 40% (29/73) | - | 1210 women (Cross sectional/7 months) | Undie 2016[57] | High | HCP training, integrated IPV-HIV consultation, assisted onsite referral contributed towards 40% uptake of referrals to on-site GBV centre |
| **Use of SRH services (n=1)** | | | | | | |
| Proportion (n) who were linked to medical care to receive lab reports on CD4 count and viral load, post-intervention | Intervention = 43.13% (69/160)<br>Control = 38.50% (30/78)<br>p = 0.493 | - | 166 women (RCT/1 month) | Brown 2018[32] | High | 7-minute integrated HIV-IPV consultation over phone had no effect on uptake of HIV services among women with experience of IPV |
| **Safety behaviour (n=2)** | | | | | | |
| Mean (SD) pre-post difference score of perceived risk and safety | Intervention = 0.33 (3.07)<br>Control = 0.13 (3.05)<br>p=0.278 | - | 166 women (RCT/1 month) | Brown 2018[32] | High | 7-minute integrated HIV-IPV consultation over phone had no effect on perceived risk and safety among HIV-positive women with experience of IPV |
| Proportion (n) who used safety plan, post-intervention | Intervention = 61.88% (99/160) | - | | | | Most HIV-positive women who received 7-minute integrated IPV-HIV consultation used safety plan and employed at least one safety strategy |
| Proportion (n) who employed at least one safety strategy | Intervention = 80% (128/160) | - | | | | |
| Proportion (n) who took an action following the IPV-enhanced HIV counselling, post-intervention | Intervention = 45.5% (25/66)<br>Control = 30.5% (18/79)<br>p=0.073 | - | 337 women (RCT/1 month) | Haberland 2016[38] | High | HCP training and ongoing support, 29-minute integrated HIV-IPV consultation, referral to IPV counsellor in ANC clinic had no effect on coping behaviour and 7 behaviour indicators of HIV care among pregnant women |
| Proportion (n) who can ask partner to use a condom, post-intervention | Intervention = 58.3% (35/107)<br>Control = 51.2% (43/134)<br>p=0.31 | - | | | | |
| **Health outcomes** | | | | | | |
| **Re-exposure to VAW (n=3)** | | | | | | |
| Proportion (n) who did not experience IPV upon partner notification of serostatus, post-intervention | Intervention = 96.9% (155/160)<br>Control = 88% (71/79) | OR = 4.37 (1.46; 13.44) | 166 HIV-positive women (RCT/1 month) | Brown 2018[32] | High | 7-minute integrated HIV-IPV consultation over phone consultation reduced IPV upon partner notification about serostatus among HIV-positive women |
| Proportion (n) who experienced any IPV, post-intervention | Intervention = 16.0% (43/337)<br>Control = 18.7% (50/351),<br>p=0.43 | - | 337 pregnant women (RCT/1 month) | Haberland 2016[38] | High | HCP training and ongoing support, 29-minute integrated HIV-IPV consultation, referral to IPV counsellor in ANC clinic had no effect on any IPV since baseline assessment |
| Mean (SD) danger assessment score | Pre-intervention = 6.02 (2.97)<br>Post-intervention = 2.82 (0.27) | MD = 3.20 (3.56) (2.43; 3.98) | 84 women (UBA/3 months) | Matseke 2013 | High | HPC training, 30-minute integrated ANC-IPV consultation, external referral contributed towards reduction in potential risk of becoming a victim of femicide among pregnant women |

| Outcomes | Results (95% CI, p) | Effect (95% CI) | N of participants (design/follow up) | Study | Risk of bias | Direction of effect/interpretation |
|---|---|---|---|---|---|---|
| **Harm (n=2)** | | | | | | |
| Proportion (n) who reported that service had not placed them in greater danger, post-intervention | Intervention = 96.25% (154/160)<br>Control = 93.67% (74/79)<br>p = 0.512 | - | 166 women (RCT/1 month) | Brown 2018[32] | High | 7-minute integrated HIV-IPV consultation over phone did not put HIV-positive women in greater danger |
| Proportion who reported harmful effects | Intervention = 0 | - | 337 pregnant women (RCT/1 month) | Haberland 2016[38] | High | HCP training and ongoing support, 29-minute integrated HIV-IPV consultation, referral to IPV counsellor in ANC clinic had no harmful effect on pregnant women |

Note. VAW violence against women. GBV gender-based violence. DV domestic violence. IPV intimate partner violence. SRH sexual and reproductive health. STI Sexually transmitted infections. HIV human immunodeficiency virus. ANC antenatal care. HCP health care provider. RCT randomised controlled trial. CBA controlled before-after study. UBA uncontrolled before-after study. CI confidence interval. SD standard deviation. IQR interquartile range. MD mean difference. OR odds ratio.
