## Supplementary material for "Interventions in sexual and reproductive health services addressing violence against women in low- and middle- income countries: a mixed-methods systematic review": The overall effect was uncertain

**Supplementary file 5. Effects and outcomes of interventions on response to VAW during routine SRH consultation plus community engagement**

| **Outcomes** | **Results** | **Effect (95% CI)** | **N of participants (design/follow up)** | **Study** | **Risk of bias** | **Direction of effect/interpretation** |
|---|---|---|---|---|---|---|
| **Direct effects on health-related cognition and emotions** | | | | | | |
| **HCP knowledge about VAW and relevant procedures (n=2)** | | | | | | |
| Proportion who knew whether there was law that deals with family violence | Pre-intervention = 71%<br>Post-intervention = 90% | 19% | ?HCPs (UBA/3 years) | Bott 2014[30] | High | Systems level activities, HCP training and ongoing support, infrastructure improvement, integrated family planning-GBV consultation, referral to onsite GBV specialist, community education contributed towards 19% increase in HCP knowledge about legal side of VAW |
| Proportion who could explain legal obligation of providers regarding family violence | Pre-intervention = 14%<br>Post-intervention = 69% | 55% | | | | |
| Proportion (n) who knew the main objectives of the programme | 100% (35/35) | | 35 HCPs (Cross sectional/4-year service data) | Sithole 2018[53] | High | HCP training, infrastructure improvement, community education on post-rape care contributed towards 25% to 100% HCP awareness about post-rape care |
| Proportion of doctors who knew the tools to monitor the programme | 25% (1/4) | | | | | |
| Proportion of doctors who knew the correct treatment guidelines | 25% (1/4) | | | | | |
| Proportion of doctors who did not know the management process | 75% (3/4) | | | | | |
| Proportion of nurses who knew the management process | 100% (27/27) | | | | | |
| HCP attitudes about VAW (n=1) | | | | | | |
| Reduction in proportion who blamed victims of physical and sexual violence (5 indicators), pre-post-intervention | - | By 29% for women provoke physical aggression<br>By 13% for men cannot control their sexual behaviour | ?HCPs (UBA/3 years) | Bott 2014[30] | High | Systems level activities, HCP training and ongoing support, infrastructure improvement, integrated family planning-GBV consultation, referral to onsite GBV specialist, community education contributed towards 13-29% reduction in negative attitudes about GBV among HCPs |
| **HCP readiness for identifying and responding to VAW (n=1)** | | | | | | |
| Reduction in proportion of cited 9 barriers to identifying IPV, pre-post intervention | - | By 29% for cultural divide between client and provider<br>By 3% for time constraints | ?HCPs (UBA/3 years) | Bott 2014[30] | High | Systems level activities, HCP training and ongoing support, infrastructure improvement, integrated family planning-GBV consultation, referral to onsite GBV specialist, community education contributed 3% to 29% reduction in perceived barriers to identifying GBV among HCPs |

| Outcomes | Results | Effect (95% CI) | N of participants (design/follow up) | Study | Risk of bias | Direction of effect/interpretation |
|---|---|---|---|---|---|---|
| Increase in proportion who felt prepared to provide counselling about emergency contraception to GBV victims | - | By 96% for counselling about emergency contraception | | | | Systems level activities, HCP training and ongoing support, infrastructure improvement, integrated family planning-GBV consultation, referral to onsite GBV specialist, community education contributed towards 95% increase in HCP preparedness to identify and respond to GBV patients |
| **Women attitudes about VAW (n=1)** | | | | | | |
| Proportion (n) who justified husband physical abuse because of childcare, post-intervention | Intervention = 41.8% (261/625)<br>Control = 45.5% (284/624) | OR = 0.81 (0.60; 1.09) | 656 women (cluster RCT/28 months) | Settergren 2018[60] | High | Systems level activities, HCP training, infrastructure improvement, integrated HIV-GBV consultation, onsite and external referral, community and couple education improved 1 out of 5 indicators of women's accepting attitudes towards VAW |
| Proportion (n) who justified husband physical abuse because she refuses to have sex with her partner, post-intervention | Intervention = 21.0% (131/625)<br>Control = 23.7% (148/624) | OR = 0.65 (0.46; 0.91) | | | | |
| Mean (SD) score of the Violence domain of the Gender Equitable Men Scale, post-intervention | Intervention = 13.17 (3.98)<br>Control = 12.51 (3.93) | MD = 1.08 (0.52; 1.65) | | | | Systems level activities, HCP training, infrastructure improvement, integrated HIV-GBV consultation, onsite and external referral, community and couple education improved women's attitudes towards more equitable gender roles |
| Mean (SD) score of the Domestic chores and daily life domain domain of the Gender Equitable Men Scale, post-intervention | Intervention = 8.74 (3.63)<br>Control = 7.62 (3.14) | MD = 1.26 (0.81; 1.71) | | | | |
| **Intermediate effects on health-related behaviour and ptractices** | | | | | | |
| **VAW enquiry rate (n=2)** | | | | | | |
| Proportion (n) who received GBV screening and counselling, post-intervention | Intervention = 88.5% (1251/1413)<br>Control = 91/7% (442/482)<br>p=0.785 | - | 656 women (cluster RCT/28 months) | Settergren 2018[60] | High | Systems level activities, HCP training, infrastructure improvement, integrated HIV-GBV consultation, onsite and external referral, community and couple education had no effect on GBV enquiry rate |
| Proportion (n) who were screened for IPV of those attended clinic | 94.5% (171/181) | - | 171 women (Cross-sectional/4 months) | Samandari 2016[49] | High | System level activities, 7-day HCP training and ongoing support, integrated family planning-IPV consultation, external referral, community education contributed towards 95% IPV enquiry rate |
| **Provision of VAW referrals (n=2)** | | | | | | |
| Proportion (n) who were referred to safe house and shelter of those screened, post-intervention | Intervention = 12.3% (173/1412)<br>Control = 2.3% (11/488)<br>p=0.216 | - | 656 women (cluster RCT/28 months) (cluster RCT/28 months) | Settergren 2018[60] | High | Systems level activities, HCP training, infrastructure improvement, integrated HIV-GBV consultation, onsite and external referral, community and couple education |

| Outcomes | Results | Effect (95% CI) | N of participants (design/follow up) | Study | Risk of bias | Direction of effect/interpretation |
|---|---|---|---|---|---|---|
| | | | | | | had no effect on rates of referrals to safe house and shelter |
| Proportion (n) who were signposted to IPV services of those disclosed | 100% (157/157) | - | 171 women (Cross-sectional/4 months) | Samandari 2016[49] | High | System level activities, 7-day HCP training and ongoing support, integrated family planning-IPV consultation, external referral, community education contributed towards 100% signposting to IPV services |
| **Provision of safety planning (n=1)** | | | | | | |
| Proportion (n) who received safety planning of those disclosed IPV | 87.3% (137/157) | - | 171 women (Cross-sectional/4 months) | Samandari 2016[49] | High | System level activities, 7-day HCP training and ongoing support, integrated family planning-IPV consultation, external referral, community education contributed towards 87% safety planning rate |
| **Provision of post-rape care (n=3)** | | | | | | |
| Mean number of rape cases presenting to hospital per month. | Pre-intervention = 8<br>Post-intervention = 13 | - | 334 survivors of sexual assault (UBA and cross-sectional/not reported) | Kim 2007[42] | High | Systems level activities, 2-day HCP training, infrastructure improvement, community education on post-rape care contributed towards increased number of rape cases presenting to hospital |
| Proportion (n) of eligible patients who received post-rape medical kit | 100% (2,081/2,081) | - | 13 sites, 2081 patients (Cross-sectional/4-year service data) | Bress 2018[31] | High | HCP training and ongoing support, infrastructure improvement, community education on post-rape care contributed towards 100% provision of post-rape care medical kit |
| Change in proportion who attended within 72 hours, over 4 years | - | 46% | 80 HCPs, 1669 patients (Cross-sectional/4 years) | Sithole 2018[53] | High | HCP training, infrastructure improvement, community education on post-rape care contributed towards improvement on 6 indicators of post-rape care provision |
| Change in proportion who received HIV post-exposure prophylaxis, over 4 years | - | 31% | | | | |
| Change in proportion who received counselling, over 4 years | - | 65% | | | | |
| Change in proportion who received HIV testing, over 4 years | - | 96.4% | | | | |
| Change in proportion who received emergency contraception, over 4 years | - | 8% | | | | |
| Change in proportion who received STI prophylaxis | - | 26% | | | | |
| **VAW disclosure rates (n=2)** | | | | | | |
| Proportion who disclosed GBV of those screened | 14% | - | ? women (UBA/3 years) | Bott 2004[30] | High | Systems level activities, HCP training and ongoing support, infrastructure improvement, integrated family planning-GBV consultation, referral to onsite GBV specialist, community education contributed towards 14% GBV identification rate |

| Outcomes | Results | Effect (95% CI) | N of participants (design/follow up) | Study | Risk of bias | Direction of effect/interpretation |
|---|---|---|---|---|---|---|
| Proportion (n) who disclosed IPV of those screened | 91.8% (157/171) | - | 171 women (Cross-sectional/4 months) | Samandari 2016[49] | High | System level activities, 7-day HCP training and ongoing support, integrated family planning-IPV consultation, external referral, community education contributed towards 92% IPV identification rate |
| Proportion (n) who disclosed GBV of those screened | 37% (49/134) | - | 134 women (Cross-sectional/5 months) | Turan 2013[56] | High | 40-hour HCP training, integrated ANC-GBV consultation, assisted external referral, community education contributed towards 37% IPV identification rate |
| **VAW referrals uptake (n=2)** | | | | | | |
| Proportion who took referral of those disclosed GBV | 30% | - | ? women (UBA/3 years) | Bott 2004[30] | High | Systems level activities, HCP training and ongoing support, infrastructure improvement, integrated family planning-GBV consultation, referral to onsite GBV specialist, community education contributed towards 30% uptake of referrals |
| Proportion (n) who took referral of those disclosed IPV | 0.6% (1/157) | - | 171 women (Cross-sectional/4 months) | Samandari 2016[49] | High | System level activities, 7-day HCP training and ongoing support, integrated family planning-IPV consultation, external referral, community education contributed towards 0.6% uptake of external referrals |
| Proportion (n) who took referral of those disclosed GBV | 53% (26/49) | - | 134 women (Cross-sectional/5 months) | Turan 2013[56] | High | 40-hour HCP training, integrated ANC-GBV consultation, assisted external referral, community education contributed towards 53% uptake of referrals |
| **Use of SRH services (n=1)** | | | | | | |
| Proportion (n) who attended any antenatal care, post-intervention | Intervention = 88.7% (1597/1800)<br>Control = 82.4% (1526/1851) | RD = 0.063 (-0.044; 0.170) | 1837 women (cluster RCT/12 months) | Cockcroft 2019[34] | Low | HCP training, infrastructure improvement, integrated DV-universal home visits that discussed domestic violence, heavy work in pregnancy, ignorance of danger signs, and lack of spousal communication with pregnant women and their spouses had no effect on 8 indicators of use of antenatal care, institutional delivery, or skilled birth attendance |
| Proportion (n) who delivered in a health facility, post-intervention | Intervention = 30.1% (475/1579)<br>Control = 21.9 (391/1785) | RD = 0.082 (-0.071; 0.235) | | | | |
| Proportion (n) who delivered by a skilled health worker post-intervention | Intervention = 29.3 (463/1579)<br>Control = 22.7 (404/1783) | RD = 0.067 (-0.081; 0.214) | | | | |
| Mean (SD of GBV client visits per facility, post-intervention | Intervention = 237.8 (110.58)<br>Control = 81.5 (46.09)<br>p=0.010 | - | 656 women (cluster RCT/28 months) | Settergren 2018[60] | High | Systems level activities, HCP training, infrastructure improvement, integrated HIV-GBV consultation, onsite and external referral, community and couple education |

| Outcomes | Results | Effect (95% CI) | N of participants (design/follow up) | Study | Risk of bias | Direction of effect/interpretation |
|---|---|---|---|---|---|---|
| | | | | | | increased use of SRH services by women with experience of GBV |
| Proportion (n) of GBV clients who received HIV testing and counselling, post-intervention | Intervention = 73.3% (1038/1416) Control = 20.9% (102/488) p=<0.001 | - | | | | Systems level activities, HCP training, infrastructure improvement, integrated HIV-GBV consultation, onsite and external referral, community and couple education increased use of 4 (HIV testing and counselling, family planning, forensic exam) out of 24 SRH services |
| Proportion (n) of GBV clients who received STI test, post-intervention | Intervention = 21.8% (308/1415) Control = 11.5% (56/488) p=0.128 | - | | | | |
| Safety behaviour (n=1) | | | | | | |
| Proportion (n) of women who used condom, post-intervention | Intervention = 17.0% (157/931) Control = 16.0% (192/1170) | PRR = 1.03 (0.85; 1.25) aPRR = 1.01 (0.84; 1.21) | 6 facilities, 1812 women (cluster RCT/35 months) | Wagman 2015[59] | High | HCP training, integrated HIV-IPV consultation, onsite referral, community education had no effect on 6 indicators of risk behaviours among women |
| **Health outcomes** | | | | | | |
| **Re-exposure to VAW (n=3)** | | | | | | |
| Proportion (n) who did not experience physical domestic violence during pregnancy, post-intervention | Intervention = 97.4 (1772/1820) Control = 90.9 (1677/1844) | RD = 0.064 (0.045; 0.084) | 1837 women (RCT/12 months) | Cockcroft 2019[34] | Low | HCP training, infrastructure improvement, integrated DV-universal home visits that discussed domestic violence, heavy work in pregnancy, ignorance of danger signs, and lack of spousal communication with pregnant women and their spouses reduced physical domestic violence |
| Proportion (n) who experienced any IPV in past 12 months, post-intervention | Intervention = 37.2% (207/556) Control = 45.7% (268/587) | OR = 0.85 (0.62; 1.16) | 6 facilities, 735 households (cluster RCT/28 months) | Settergren 2018[60] | High | Systems level activities, HCP training, infrastructure improvement, integrated HIV-GBV consultation, onsite and external referral, community and couple education had no effect on any IPV |
| Proportion (n) who experienced physical IPV in past 12 months, post-intervention | Intervention = 12% (217/1812) Control = 16% (346/2127) | PRR = 0.74 (0.63; 0.86) aPRR = 0.79 (0.67; 0.92) | 6 facilities, 1812 women (cluster RCT/35 months) | Wagman 2015[59] | High | HCP training, integrated HIV-IPV consultation, onsite referral, community education reduced incidents of physical and sexual IPV and had no effect on emotional IPV. |
| Proportion (n) who experienced emotional IPV in past 12 months, post-intervention | Intervention = 18% (311/1737) Control = 20% (409/2039) | PRR = 0.89 (0.78; 1.02) aPRR = 0.91 (0.79; 1.04) | | | | |
| Proportion (n) who experienced sexual IPV in past 12 months, post-intervention | Intervention = 10% (167/1737) Control = 13% (261/2038) | PRR = 0.75 (0.62; 0.90) aPRR = 0.80 (0.67; 0.97) | | | | |
| **Sexual and reproductive health (n=1)** | | | | | | |

| Outcomes | Results | Effect (95% CI) | N of participants (design/follow up) | Study | Risk of bias | Direction of effect/interpretation |
|---|---|---|---|---|---|---|
| Proportion (n) who did not have swelling of face or hands, post-intervention | Intervention = 97.4% (1790/1837)<br>Control = 71.1% (1317/1853) | RD = 0.264 (0.194; 0.333) | 1837women (RCT/12 months) | Cockcroft 2019[34] | Low | HCP training, infrastructure improvement, integrated DV-universal home visits that discussed domestic violence, heavy work in pregnancy, ignorance of danger signs, and lack of spousal communication with pregnant women and their spouses improved 9 out of 13 indicators of pregnancy and postpartum complications |
| Proportion (n) who did not have raised blood pressure, post-intervention | Intervention = 96.6% (1409/1458)<br>Control = 85.1% (1269/1492) | RD = 0.116 (0.042; 0.190) | | | | |
| Proportion (n) who did not have post-partum sepsis, post-intervention | Intervention = 81.1% (1478/1822)<br>Control = 48.8% (903/1852) | RD = 0.324 (95% CI 0.115; 0.493) | | | | |
| Incidence of HIV per100 person-years | Intervention = 0.99<br>Control = 1.15 | IRR = 0.86 (0.61; 1.22<br>aIRR = 0.72 (0.49; 1.07)) | 6 facilities, 1812 women (cluster RCT/35 months) | Wagman 2015[59] | High | HCP training, integrated HIV-IPV consultation, onsite referral, community education had no effect on incidence of HIV among women |

Note. VAW violence against women. GBV gender-based violence. DV domestic violence. IPV intimate partner violence. SRH sexual and reproductive health. STI Sexually transmitted infections. HIV human immunodeficiency virus. ANC antenatal care. HCP health care provider. RCT randomised controlled trial. CBA controlled before-after study. UBA uncontrolled before-after study. CI confidence interval. SD standard deviation. IQR interquartile range. MD mean difference. OR odds ratio. RD risk difference. IRR incidence rate ratio.
