## Supplementary material for "Interventions in sexual and reproductive health services addressing violence against women in low- and middle- income countries: a mixed-methods systematic review": This intervention category had the most robust evidence from six RCTs and one UBA study

### Supplementary file 6. Effects and outcomes of interventions on response to VAW in addition to routine SRH consultation

| Outcomes | Results | Effect (95% CI) | N of participants (design/follow up) | Study | Risk of bias | Direction of effect/interpretation |
|---|---|---|---|---|---|---|
| **Direct effects on health-related cognition and emotions** | | | | | | |
| **Women knowledge about VAW (n=1)** | | | | | | |
| Proportion who recognised violence as an issue of power, post-intervention | 60.6% | | 155 women (UBA/ First prenatal appointment to 6 weeks after delivery) | Arora 2019[29] | High | After 2 or more 30-45-minute psychosocial counselling sessions, around 60% of pregnant women were aware about domestic violence and its impact on health |
| Proportion who recognised the impact of violence on health, post-intervention | 65.5% | | | | | |
| **Women readiness for addressing VAW (n=1)** | | | | | | |
| Proportion who recognised the need to take steps to address violence, post intervention | 59.9% | | 155 women (UBA/ First prenatal appointment to 6 weeks after delivery) | Arora 2019[29] | High | After 2 or more 30-45-minute psychosocial counselling sessions, 60% of pregnant women were ready to address VAW |
| **Intermediate effects on health-related behaviour and practices** | | | | | | |
| **VAW referral uptake (n=1)** | | | | | | |
| Proportion (n) who used specialist IPV services, post-intervention | Intervention = 0.96% (1/104)<br>Control = 1.00% (1/100) | - | 110 women (RCT/Prenatal appointment to 1 week after delivery) | Cripe 2010[35] | Some concerns | 1 30-minute psychosocial counselling session, resource card, external referral had no effect on uptake of external referrals |
| **Use of SRH services (n=1)** | | | | | | |
| Proportion (n) who missed antiretroviral medication, post-intervention | Intervention = 42.3% (19)<br>Control = 36.4% (25) | - | 32 women (RCT/6 months) | Sikkema 2018[52] | High | 7 90-minute psychosocial sessions on coping had no effect on engagement with HIV treatment among women with a history of sexual violence |
| Proportion (n) with high levels of non-retention in care, post-intervention | Intervention = 42.3% (26)<br>Control = 33.3% (27) | - | | | | |
| **Use of non-health services (n=1)** | | | | | | |
| Proportion (n) who used legal services, post-intervention | Intervention = 1.92% (2/104)<br>Control = 3.00% (3/100) | - | 110 women (RCT/Prenatal appointment to 1 week after delivery) | Cripe 2010[35] | Some concerns | 1 30-minute psychosocial counselling session, resource card, external referral had no effect on use of community resources among pregnant women |
| Proportion (n) who used police, post-intervention | Intervention = 0.96% (1/104)<br>Control = 4.00% (4/100) | - | | | | |
| Proportion (n) who used social services, post-intervention | Intervention = 1.92% (2/104)<br>Control = 2.00% (2/100) | - | | | | |
| **Safety behaviour (n=4)** | | | | | | |
| Mean (SD) score of using safety behaviours, post-intervention | Intervention = 9.50 (2.63)<br>Control = 7.74 (2.42) | MD = 2.41 (1.43; 3.40) | 70 women (RCT/Prenatal appointment to 6 weeks after delivery) | Sapkota 2020[50] | Low | 1 35-45-minute psychosocial counselling session, resource card, contact details of the counsellor increased use of safety behaviours among pregnant women |

| Outcomes | Results | Effect (95% CI) | N of participants (design/follow up) | Study | Risk of bias | Direction of effect/interpretation |
|---|---|---|---|---|---|---|
| Proportion who adopted safety behaviours, post-intervention | Intervention = 30.3%<br>Control = 11.2% | | 110 women (RCT/Prenatal appointment to 1 week after delivery) | Cripe 2010[35] | Some concerns | 1 30-minute psychosocial counselling session, resource card, external referral had no effect on safety behaviours among pregnant women |
| Mean (SD) score of avoidance, coping post-intervention | Intervention = 2.17 (0.13)<br>Control = 1.99 (0.09) | - | 32 women (RCT/6 months) | Sikkema 2018[52] | High | 7 90-minute psychosocial training sessions reduced avoidance coping, but had no effect on social coping among women with a history of sexual violence |
| Mean (SD) score of social coping, post-intervention | Intervention = 2.90 (0.10)<br>Control = 2.58 (0.10) | - | | | | |
| Proportion (n) who used adaptive coping strategies at individual level pre- and post-intervention | Pre-intervention = 51.4% (73)<br>Post-intervention = 59.1% (84)<br>p=0.193 | | 155 women (UBA/ First prenatal appointment to 6 weeks after delivery) | Arora 2019[29] | High | 2 or more 30-45-minute psychosocial counselling sessions had no effect on coping behaviours among pregnant women |
| Proportion who used adaptive coping strategies at informal and formal levels pre- and post-intervention | Pre-intervention = 85.2% (121)<br>Post-intervention = 86.6% (123)<br>p=0.832 | | | | | |
| **Health outcomes** | | | | | | |
| **Re-exposure to VAW (n=4)** | | | | | | |
| Mean (SD) score of verbal and physical IPV, post-intervention | Intervention = 11.62 (2.05)<br>Control = 13.28 (1.94)<br>p<0.001 | - | 50 pregnant women (RCT/2 months) | Khalili 2019[40] | High | 4 90-minute psychoeducational counselling sessions reduced verbal and physical IPV among pregnant women |
| Mean (SD) score of total IPV, post-intervention | Intervention = 17.70 (11.12)<br>Control = 31.22 (21.17) | MD = 13.51 (9.99; 17.02) | 141 pregnant women (RCT/6 months) | Mutisya 2018[47] | High | 1-3 30-35-minute psychosocial counselling sessions, risk assessment, safety planning, resource card, external referral reduced IPV among pregnant women |
| Proportion who experienced physical IPV, post intervention | Intervention = 51.2%<br>Control = 65.9% | RR = 0.78 (0.63; 0.93) | 125 women (RCT/3 months) | Tanghizaden 2018[55] | Some concerns | 4 90-minute psychosocial training sessions on problem-solving skills reduced physical and psychological IPV but had no effect on sexual IPV among pregnant women |
| Proportion who experienced psychological IPV, post intervention | Intervention = 67.4%<br>Control = 92.4% | RR = 0.73 (0.64; 0.83) | | | | |
| Proportion who experienced sexual IPV, post intervention | Intervention = 50.4%<br>Control = 57.6% | RR = 0.87 (0.69; 1.09) | | | | |
| Change in proportion who experienced physical domestic violence, before-after | 74.6% to 3.5% | - | 155 women (UBA/ First prenatal appointment to 6 weeks after delivery) | Arora 2019[29] | High | 2 or more 30-45-minute psychosocial counselling sessions reduced physical, emotional, and financial domestic violence among pregnant women |

| Outcomes | Results | Effect (95% CI) | N of participants (design/follow up) | Study | Risk of bias | Direction of effect/interpretation |
|---|---|---|---|---|---|---|
| Change in proportion who experienced emotional domestic violence, before-after | 98.6% to 34.5% | - | | | | |
| Change in proportion who experienced financial domestic violence, before-after | 72.5% to 11.3% | - | | | | |
| **Mental health (n=6)** | | | | | | |
| Mean (SD) score of anxiety, post-intervention | Intervention = 4.33 (3.84)<br>Control = 6.93 (4.87) | MD = -3.73 (-5.42; -2.04) | 70 women (RCT/prenatal appointment to 6 weeks after delivery) | Sapkota 2020[50] | Low | 1 35-45-minute psychosocial counselling session, resource card, contact details of the counsellor reduced anxiety and depression among pregnant women |
| Mean (SD) score of depression, post-intervention | Intervention = 3.51 (3.46)<br>Control = 6.13 (3.68) | MD = -3.41 (-4.84; -1.99) | | | | |
| Mean (SD) score of postnatal depression, post-intervention | Intervention = 5.34 (4.23)<br>Control = 12.46 (4.22) | MD = 7.12 (6.21; 8.03) | 141 women (RCT/6 months) | Mutisya 2018[47] | High | 1-3 30-35-minute psychosocial counselling sessions with risk assessment, safety planning, external referral, and resource card reduced depression among pregnant women |
| Mean (SD) score of PTSD, post-intervention | Intervention = 28.61 (5.04)<br>Control = 22.50 (3.47) | - | 32 women (RCT/6 months) | Sikkema 2018[52] | High | 7 90-minute psychosocial training sessions had no effect on PTSD symptoms among women with a history of sexual violence |
| Mean (SD) score of psychological distress, post-intervention | Intervention = 22.28 (3.81)<br>Control = 24.06 (4.16)<br>p<0.001 | - | 50 women (RCT/2 months) | Khalili 2019[40] | High | 4 90-minute psychoeducational counselling sessions reduced psychological distress among pregnant women |
| Difference between baseline and post-intervention mean (SD) score for mental health | Intervention = 2.50 (20.95)<br>Control = 2.04 (19.61) | MD = 4.54 (-1.07; 10.15) | 110 women (RCT/Prenatal appointment to 1 week after delivery) | Cripe 2010[35] | Some concerns | 1 30-minute psychosocial counselling session, resource card, external referral had no effect on mental health of pregnant women |
| Change in proportion who experienced any emotional health problems | 96.5% to 33.1% | - | 155 women (UBA/ First prenatal appointment to 6 weeks after delivery) | Arora 2019[29] | High | 2 or more 30-45-minute psychosocial counselling sessions reduced % of pregnant women with emotional health problems. |
| **Sexual and reproductive health (n=1)** | | | | | | |
| Proportion (n) of those with unsuppressed HIV viral load, post-intervention | Intervention = 15.8% (19)<br>Control = 20.0% (25)<br>($\chi2$ (1) = 0.13, p = 0.72) | - | 32 women (RCT/6 months) | Sikkema 2018[52] | High | 7 90-minute psychosocial training sessions had no effect on adherence to therapy measured by HIV viral load among women with a history of sexual abuse |
| **Physical health (n=2)** | | | | | | |
| Difference between baseline and post-intervention mean (SD) score for general health | Intervention = 5.30 (15.62)<br>Control = 4.74 (14.67) | MD = 0.05 (-6.80; 7.79) | 110 women (RCT/Prenatal appointment to 1 week after delivery) | Cripe 2010[35] | Some concerns | 1 30-minute psychosocial counselling session, resource card, external referral had no effect on general health |

| Outcomes | Results | Effect (95% CI) | N of participants (design/follow up) | Study | Risk of bias | Direction of effect/interpretation |
|---|---|---|---|---|---|---|
| Change in proportion who experienced any physical health problems | 54.6% to 10.5% | - | 155 women (UBA/ First prenatal appointment to 6 weeks after delivery) | Arora 2019[29] | High | 2 or more individual psychosocial counselling sessions reduced % of pregnant women with physical health problems. |
| **Quality of life (n=2)** | | | | | | |
| Mean (SD) score of overall quality of life, post-intervention | Intervention = 17.22 (3.00)<br>Control = 15.19 (2.77) | MD = 2.45 (1.51; 3.39) | 70 women (RCT/prenatal appointment to 6 weeks after delivery) | Sapkota 2020[50] | Low | 1 35-45-minute psychosocial counselling session, resource card, contact details of the counsellor improved quality of life among pregnant women |
| Difference between baseline and post-intervention mean (SD) score for physical functioning | Intervention = -15.67 (28.35)<br>Control = -15.70 (25.06) | MD = 0.03 (-7.37; 7.42) | 110 women (RCT/Prenatal appointment to 1 week after delivery) | Cripe et 2010[35] | Some concerns | 1 30-minute psychosocial counselling session, resource card, external referral had no effect on general health, bodily pain, vitality, social functioning among pregnant women. |
| Difference between baseline and post-intervention mean (SD) score for bodily pain | Intervention = -7.40 (28.33)<br>Control = -7.90 (24.28) | MD = 0.50 (-6.80; 7.79) | | | | |
| Difference between baseline and post-intervention mean (SD) score for vitality | Intervention = -0.19 (22.34)<br>Control = -3.65 (22.06) | MD = 3.46 (-2.67; 9.59) | | | | |
| Difference between baseline and post-intervention mean (SD) score for social functioning | Intervention = -0.36 (34.94)<br>Control = -3.50 (37.06) | MD = 3.14 (-6.80; 13.08) | | | | |

Note. VAW violence against women. GBV gender-based violence. DV domestic violence. IPV intimate partner violence. SRH sexual and reproductive health. STI Sexually transmitted infections. HIV human immunodeficiency virus. ANC antenatal care. HCP health care provider. RCT randomised controlled trial. CBA controlled before-after study. UBA uncontrolled before-after study. CI confidence interval. SD standard deviation. IQR interquartile range. MD mean difference. RR relative risk.
