## Supplementary file 7 summarises factors that women and HCPs perceived as barriers to intervention implementation and impact. for "Interventions in sexual and reproductive health services addressing violence against women in low- and middle- income countries: a mixed-methods systematic review"

### Supplementary file 7. Barriers to identification and response to VAW in sexual and reproductive health services

| Theme | Description | Discussed by | Supporting text |
|---|---|---|---|
| Acceptability of VAW | Attitudes and social norms that regulate the acceptability of VAW at individual and community levels | Cristofides 2010[33]<br>Laisser 2011[45]<br>Smith 2013[54]<br>Turan 2013[56] | **Women:** "Women did not want a referral because they did not feel the violence was serious or they felt these were personal issues that they will solve on their own"[38]<br>**HCPs:** "Some of these patients are themselves to be blamed. You know some women don't want to be polite to their husbands and adhere to the norms in their marriages that is why they are beaten. It takes time, need to be more patient and expertize to screen, which we miss. It may be too costly for training. (FGD3 Male Nurse)"[45]<br>**Community:** "So when somebody is saying that women are not supposed to be beaten, that. . . they should go to somebody and take some action, in the community it is like that person is acting against the will of the community. To the men it is like he is an outcast in the community, an outlaw who is not supposed to be there. . . . In social places you will hear them saying that he is not a good person because if he is preaching to our ladies and women to take action against us, then it is like he wants to bring a revolution, women are going to overpower us and then we are going to be voiceless. (Focus Group #1, Respondent #8)"[56] |
| Fear of negative consequences | Real or potential negative consequences (psychological, legal, financial) of engaging in VAW work that could make the situation worse for individuals and health system | Bott 2004[30]<br>Christofides 2010[33]<br>Haberland 2016[38]<br>Knettel 2019[44]<br>Laisser 2011[45]<br>Samandari 2016[49]<br>Sapkota 2020[51]<br>Sithole 2018[53] | **Women:** "The sessions would irritate me when we talked about my rape; I hated to talk about it even though when I had talked about it, I would feel better. My heart would feel sore. Even talking about my HIV status irritated me because I still beat myself for infecting my child"[44]<br>"a few women stated that privacy concerns made it difficult for them to participate in the intervention, especially the group sessions. One participant explained that she was "afraid that I might be seen by a participant who knows me and who might go around discussing my problems."[44]<br>**HCPs:** "Providers were responsible for all IPV screening and counseling, as well as their regular FP duties. This led not only to an increased burden of duty for providers, but also the experience of secondary trauma, resulting from the exposure to clients' IPV stories."[49]<br>**Healthcare system**: "In one shift we normally attend up to 60 plus in a room for the two clinicians. Sometimes we reach up to 100 clients when it is a busy day, but if we are to attend one client at a time then it will be only 15 clients per day in a room. Where will others go?" Male clinician"[45] |
| Limited readiness for VAW work in SRH services | Structural unreadiness within health system: lack of support from leadership, time pressure, insufficient budget, lack of adequate resources, limited privacy | Abeid 2016[28]<br>Bott 2004[30]<br>Christofides 2010[33]<br>Haberland 2016[38]<br>Laisser 2011[45]<br>Sammandari 2016[49]<br>Undie 2016[57] | "System level factors may have influenced the implementation of IPV screening by lay counsellors …Other factors included inadequate management and supervision, burn-out, and small stipends which adversely affect counsellors' motivation to do something perceived as extra."[33]<br>"The HCWs felt they had not much to offer to the women who were experiencing IPV. This category thus represents an uncertainty as to whether the health care system is ready for routine screening for IPV and suggests a need for reinforced organizational change."[45]<br>"Perceived barriers to replication and scale-up included inadequate funding, insufficient clinic staff, and lack of political commitment for IPV services on the part of MOHPH"[49]<br>"Providers' main criticism was the longer time required to conduct the enhanced counseling. It created delays in the system, frustrating clients who were tired of being at the hospital for so long. Providers also felt the effects of extra time. One noted explicitly that they are supposed to see a certain number of clients each day and if they do not meet their targets they will have problems with management."[38]<br>"D3: It is also difficult to examine a patient in front of another one even if we use curtains. There is one examination bed for two of us and when you ask questions about STD patients feel embarrassed. Although we try to use low voice, people like to listen to others' conversations. (FGD1 Female Clinician"[45] |
| | Wider systems unreadiness: lack of services to refer to, poor referral system, untrained staff in non-health services | Abeid 2016[28]<br>Bott 2004[30]<br>Christofides 2010[33]<br>Laisser 2011[45] | "Providing referrals to women who disclosed current experiences of IPV may be of limited utility where services are hard to access"[33] |
| | Society unreadiness: poverty, no money for transport fare and | Haberland 2016[38]<br>Knettel 2019[44]<br>Laisser 2011[45] | "D1: You know I have nothing much to say but would like to do the screening -the resources are my dilemma. Many women are poor 'wanyonge' and are not strong enough to fight with their husbands but maybe this would be their good start. They will be happier later in future. (FGD3 Female Clinician)"[45] |

| | | | |
|---|---|---|---|
| | healthcare services, no transport, financial dependence on husband. | Sithole 2018[53] | "Her issue was that the partner used to beat her and to abuse her physically when she asked for bus fare to come to the clinic. So you see at the end of the day if she doesn't get help to deal with the violence she won't be able to come to the clinic because she is being abused when she asks for money to come to the clinic and it will affect her overall outcome…"[38] "Getting to the clinic would be a challenge as I do not work and often had to borrow money."[44] |
| | Women's unreadiness for VAW services offered by HCPs: the demand-supply gap between women's preferences for adequate response to VAW and what HCPs offered to them | Christofides 2010[33] Haberland 2016[38] Undie 2016[57] | "However, one woman questioned whether there was any point in talking to the lay counselor unless the counselor would go home and make her husband stop. Other participants, who had not disclosed abuse, suggested that health care providers could talk to a woman's abusive partner and this would stop the violence. Others suggested that if abusive partners knew about IPV screening they would stop. This seems perhaps unrealistic."[33] |
| | | | "Although the initial intention of the intervention was for IPV-positive clients to receive same-day services at the GBV clinic, this was not always possible, because women did not have the time, GBV clinic staff were not always available, and some clients preferred to have their initial GBV clinic appointment on a later date. There were occasions when providers referred clients to the GBV clinic and the clients initially complied with the referral, only to find that their needs could not be attended to immediately due to the unavailability of staff atthe GBV clinic."[57] |
